# A Continuously Active Antimicrobial Coating effective against Human Coronavirus 229E

**DOI:** 10.1101/2020.05.10.20097329

**Authors:** Luisa A. Ikner, Jason R. Torrey, Patricia M. Gundy, Charles P. Gerba

## Abstract

The disinfection of high-contact surfaces is seen as an infection control practice to prevent the spread of pathogens by fomites. Unfortunately, recontamination of these surfaces can occur any time after the use of common disinfectants. We recently reported on a novel continuously active antimicrobial coating which was shown to reduce the spread of healthcare acquired infections in hospitals. We evaluated a modified coating that demonstrated a residual efficacy against viruses. The coated surfaces were found to be effective against human coronavirus (HCoV) 229E, reducing the concentration of these viruses by greater than 90% in 10 minutes and by greater than 99.9% after two hours of contact. The coating formulation when tested in suspension yielded a greater than 99.99% reduction of HCoV 229E within ten minutes of contact. This outcome presents an opportunity for controlling the transmission of COVID-19 from contaminated fomites.

---

Fomites are inanimate surfaces that may harbor both enteric and respiratory human pathogenic viruses (Boone and Gerba, 2007). Surface disinfection reduces levels of infectious viruses, thereby lowering the potential for their spread in commercial and healthcare facilities, as well as residential environments (Sifuentes et al, 2014; Beamer et al, 2015; Sassi et al, 2015; Reynolds et al 2016, 2019). Unfortunately, recontamination of these surfaces can occur at any time following the use of common liquid disinfectants that are generally wiped dry from surfaces following treatment. Several self-disinfecting surfaces capable of inactivating deposited pathogens have been developed during recent years (Querido et al, 2019). We recently reported on a novel continuously active antimicrobial coating that was shown to reduce the spread of hospital acquired infections in clinical settings (Ellingson et al, 2020; Tamimi et al, 2014). The manufacturer recently developed a second generation of this technology that was designed to provide additional anti-viral action. The purpose of this study was to assess the efficacy of this technology against a coronavirus. Coronavirus 229E, one of the agents of the common cold was used in this study.

## Methods

### Sample preparation

The antimicrobial coated surfaces with SurfaceWise2™ (a quaternary ammonium polymer coating) were kindly prepared and supplied by Allied BioScience, Inc (Dallas, Texas). Stainless steel coupons (annealed 304) measuring 2” x 2” were cleaned with acetone, sterilized by autoclaving, and arranged on a panel for spraying. A robotic slide was equipped with an electrostatic sprayer, with coating coverage monitored by an XRF (X-Ray Fluorescence Spectroscopy) spectrometer analyzer to ensure application of a uniform coating. After the desired coating thickness was reached, the coupons were removed from the panel and the coating was cured overnight. Evenness of coating coverage was again confirmed using a hand-held XRF spectrometer.

### Preparation of the test virus

Human coronavirus 229E (HCoV) (ATCC VR-740), an enveloped respiratory virus, was propagated and assayed in the human lung fibroblast MRC-5 cell line (ATCC CCL-171). Infected cells were subjected to three freeze-thaw cycles after cytopathogenic effects (CPE) were observed in the monolayer to release the viruses, and the cell lysates were centrifuged at 1000 x g for 10 minutes to pellet the cell debris for disposal. The viruses in the supernatant then underwent a polyethylene glycol [9% (w/v), MW 8000] extraction with 0.5 M sodium chloride, with mixing overnight at 4 °C. After centrifugation at 10,000 x g for 30 min, the pelleted virus was resuspended in 0.01 M phosphate buffered saline (PBS; pH 7.4) (Sigma, St. Louis, MO) to approximately 5% of the original virus suspension volume. The HCoV 229E stocks were then aliquoted and stored at –80°C.

HCoV 229E viral stocks were enumerated on MRC-5 cells seeded into 96-well cell culture trays using the TCID_50_ (tissue culture infectious dose at the 50% endpoint) technique as described by Payment and Trudel (1993). This technique determines the dilution at which 50% of the wells show cytopathogenic effects. Taking the inverse log of this dilution gives a titer of the virus per ml TCID_50_. The minimum detection for the method described herein was 3.7 viruses per ml.

### Test procedures

The American Society of Testing and Materials (ASTM) Method E1153 was used to assess the performance of the anti-viral coating. ASTM E1153 is a quantitative method used to evaluate the efficacy of sanitizers on pre-cleaned inanimate, nonporous, non-food contact surfaces. Per the method, products are evaluated against a representative Gram-negative and Gram-positive organism with a maximum contact time of 5 minutes. This method was modified for use in the current study to assess the efficacy of SurfaceWise2 applied to stainless steel coupons against human coronavirus 229E. Coated stainless steel coupons were held under ambient conditions for 14 days after treatment. On the day of testing, 0.1 mL of viral suspension (no soil load) was inoculated onto the coated stainless steel carriers and incubated for a contact time of 120 minutes at room temperature. At conclusion of the contact time, a cotton-tipped swab pre-dipped in Letheen Broth Base (LBB) (Hardy Diagnostics, Santa Martin, CA) was used to neutralize the test material and to extract remaining infectious viruses from the test surface. The swab was then placed into a tube containing 1 mL of LBB, and vortexed vigorously to release the viruses. The entirety of the virus suspension was immediately passed through a Sephadex G-10 (Sigma, St. Louis, MO) gel filtration column for secondary neutralization, and to decrease cytotoxic effects on MRC-5 host cell monolayers. Columns were centrifuged for 5 minutes at 4,000 x g to extract the liquid. Appropriate 1:10 dilutions of neutralized control and test extracts were made using Eagle Minimal Essential Media (MEM) (Mediatech, Manassas, CA), followed by plating onto MRC-5 monolayers prepared in 96-well trays in replicates of six per dilution (0.050 mL per well). The efficacy of the test substance was determined by comparing the amount of reduction in virus titer between the control and test conditions and calculating the log_10_ reduction. The test procedure is shown in Figure 1.

**Figure 1.**
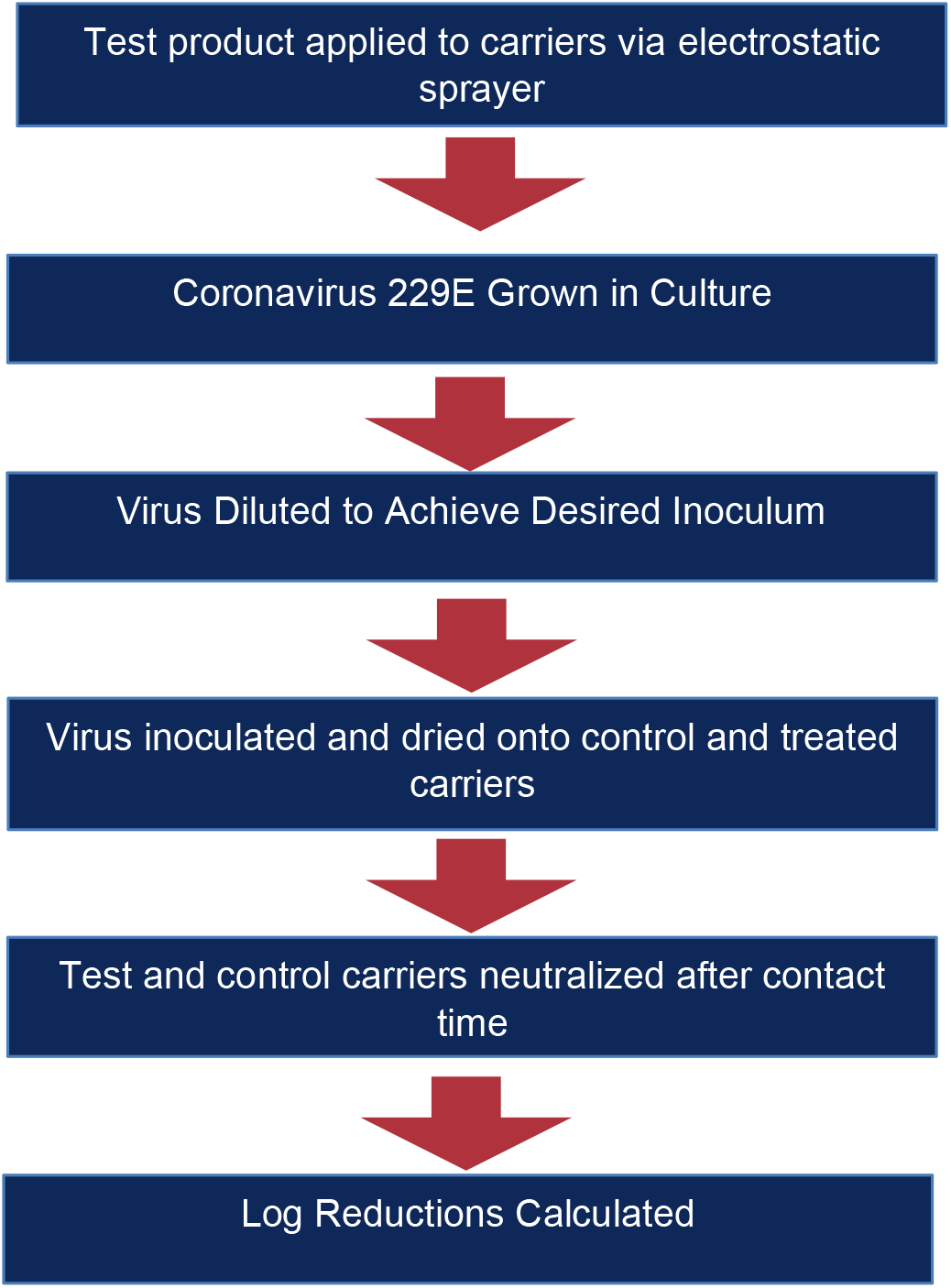
Diagram of the procedure.

Antiviral efficacy of SurfaceWise 2 was further evaluated in liquid form using the ASTM E1052 Standard Test Method. The test virus was prepared as described previously, and 0.1 mL of the virus stock suspension were added to 0.9 mL of the test substance (Ready-to-Use liquid) followed by a brief vortex and contact time of 10 minutes at room temperature. A separate tube containing 0.9 mL of MEM was used as a control. At conclusion of the contact time, 0.1 mL of test suspension was added to 0.9 mL Letheen Base Neutralizing Broth (Hardy Diagnostics, Santa Martin, CA). After brief vortexing, the total volume of the neutralized test suspension was passed through a Sephadex G-10 gel filtration column followed by gel filtration as previously described. Appropriate dilutions (1:10) of neutralized control and test conditions were made in MEM and plated in replicates of four per dilution on MRC-5 cell monolayers prepared in 24-well trays (0.1 mL pre replicate well). Viral titers in the Control and Test Suspensions were calculated using the Spearman-Karber Method. The efficacy of the test substance is determined by calculating the log_10_ reduction of infectious viruses remaining after exposure to the test product for the 10-minute contact time relative to the infectious viral titer in the control suspension.

## Results

The results shown in Table 1 demonstrate a 1.34 log (>90 %) reduction of the virus in ten minutes compared to the control untreated surface and a greater that 3.99 logs (>99.99%) within two hours.

**Table 1.** Inactivation of human coronavirus 229E on SurfaceWise2-treated vs. nontreated stainless steel (SS) surfaces.

| Surface Type | Contact Time (minutes) | Recovered Viral Titer (TCID<sub>50</sub> / ml) | Mean Recovered Viral Titer (TCID<sub>50</sub> / ml) | Log<sub>10</sub> Reduction |
| --- | --- | --- | --- | --- |
| Untreated SS Control | 10 | 9.28E+04<br>4.31E+04<br>2.94E+04 | 5.51E+04 | N/A |
| SurfaceWise2 SS | 10 | 2.94E+02<br>2.94E+03<br>4.31E+03 | 2.51E+03 | 1.34 |
| Untreated SS Control | 120 | 6.32E+04<br>2.94E+04<br>9.28E+04 | 6.18E+04 | N/A |
| Treated | 120 | <6.32<br><6.32<br><6.32 | <6.32 | >3.99 |

NA = not applicable

Table 2 also demonstrate that liquid form of ready-to-use SurfaceWise™ reduced coronavirus titer by >4.7 log_10_ (>99.99%).

**Table 2.** Inactivation of human coronavirus 229E in suspension by SurfaceWise2™.

2. Inactivation of human coronavirus 229E in suspension by SurfaceWise2™
| Suspension | Contact Time (minutes) | Viral Titer (TCID<sub>50</sub>/ml) | Mean Viral Titer (TCID<sub>50</sub>/ml) | Log Reduction |
| --- | --- | --- | --- | --- |
| MEM (Control) | 10 | 1.78E+06<br>1.78E+06 | 1.75E+06 | N/A |
| Antimicrobial Ready-to-use | 10 | < 3.16E+01<br>< 3.16E+01 | < 3.16E+01 | > 4.75<br>> 4.75 |

## Discussion

Antiviral coatings offer the advantage of rendering viruses noninfective following contact with a treated surface. Antimicrobial-coated surfaces are not meant to substitute for regular cleaning and disinfection of surfaces, but rather offer an additional barrier to reduce human exposure to infectious viruses from fomites. During the fall season, respiratory viruses such as influenza (Boone and Gerba, 2005) and parainfluenza (Boone and Gerba, 2010) can be found on third or more of common high-touch fomites, and therefore serve as an exposure route for the transmission of these and other infectious agents. It has been found that the contamination of a push plate door entrance into an office building can lead to contamination of 50% of the commonly-touched surfaces and hands of office workers within four hours (Reynolds et al, 2016). Interventions that employ disinfecting wipes have been shown to reduce the probability of infection in office settings (Beamer et al, 2015). Antimicrobial coatings could provide an additional novel mean of protection in reducing the spread of coronaviruses in indoor environments and public places where there is continuous contamination.

## Data Availability

The data and calculations for each experiment can be provided

